# Prevalence and predictors of self-medication for COVID-19 among slum dwellers in Jinja City, Uganda

**DOI:** 10.1101/2023.09.08.23295267

**Authors:** Prossy Nakito, Angela N. Kisakye, Abel Wilson Walekhwa, Gloria Tumukunde, Charity Mutesi, Nicholas Muhumuza, Carolyne Nyamor, David Musoke, Geofrey Musinguzi, Dathan M. Byonanebye

**Affiliations:** Department of Community Health and Behavioural Sciences, School of Public Health, Makerere University, Kampala, Uganda; African Field Epidemiology Network Lugogo House, Plot 42 Lugogo By-Pass, Kampala; Makerere University School of Public Health, Department of Health Policy Planning and Management; Diseases Dynamics Unit, Department of Veterinary Medicine, University of Cambridge, United Kingdom; Department of Disease Control and Environmental Health, University School of Public Health; Kirby Institute, University of New South Wales, Sydney, Australia

**Keywords:** Self-medication, COVID-19, Slum dwellers, Confirmed COVID-19, Self-reported COVID-19

## Abstract

**Introduction:** Self-medication is a serious public health concern globally and is more prevalent in underserved populations, especially in resource limited settings. The lack of effective treatment for COVID-19 and poor access to healthcare were drivers of self-medication. We investigated the prevalence and associated factors with self-medication for COVID-19 among slum dwellers in a Ugandan slum.

**Methods and materials:** We conducted a cross-sectional study using randomly selected respondents from slums in Jinja city, Uganda. Households were proportionately selected from the slums and one participant with confirmed or self-reported COVID-19 during 2021 was recruited. Poisson regression with robust standard errors was used to determine the crude (CPR) and adjusted prevalence ratios (APR) (95% CI) of factors associated with self-medication. Variables were selected apriori and backward elimination approach used to fit the final multivariate model in which variables with a *P*≤ 0.05 were included.

**Results:** Overall, 517 respondents were recruited, median age (years) was 31 (26-40), and 59% were male. The prevalence of self-medication for COVID-19 was 87.23% (451/517), 95% CI: [84.00%-90.00%] and 56% knew that self-medication was dangerous. Age≥50 years, compared to 20-29 years [APR: 1.12, 95% CI:1.05, 1.20], being female [APR: 1.07, 95% CI: 1.02, 1.13], minor [APR: 1.62, 95% CI: 1.25, 2.11], and severe symptoms [APR: 1.51, 95% CI: 1.16, 1.96], access to internet [APR: 1.13, 95% CI: 1.07, 1.20]. Having medical insurance [APR: 0.63, 95% CI: 0.46, 0.87] and awareness about laws against self-medication [APR: 0.89, 95% CI: 0.81, 0.97] were associated with a lower risk of self-medication.

**Conclusion:** The prevalence of self-medication in slum dwellers in Uganda was high despite high awareness about its dangers. Self-medication was common in those with severe symptoms and those access to internet. There is need to control infodemia and improve health insurance cover in informal settlements within Uganda.

## Introduction

While Africa did not registered very high incidence of Coronavirus disease 2019 (COVID-19) compared to the rest of the word, the disease had significant impact on people living in informal settlements. As of February 2023, the World Health Organization (WHO) estimated that there were more than 9.4 million cases of COVID-19, with approximately 173,295 deaths [1, 2]. Uganda experienced a first wave and a more serious second wave between November to December 2020 and April to June 2021, respectively [3]. The second wave of the pandemic was fuelled by five different variants and a high mortality rate, estimated at 36% [4]. Cumulatively, the country had reported 170,409 confirmed cases and 3,630 deaths by February 28^th^, 2023 [5]. Amidst this burden, effective treatments for COVID-19 were not accessible in Africa, and the government treatment guidelines focused on symptom management [2].

COVID-19 impacted nearly every sector of development the world over. The health sector was not spared and it experienced monumental challenges in trying to cope with and respond to the pandemic [6]. Several countries instituted strategies such as restrictions on travel to reduce disease transmission and the burden on the health [7, 8]. However, in resource limited settings, these measures exacerbated the existing challenge of limited access to health services, especially in underserved communities. The infodemia around COVID-19 infection, overstretched health systems and limitations in access to health services exacerbated self-medication [9]. The World Health Organization (WHO) identified self-medication as a significant public health issue with risks if not used properly [10]. It defines self-medication as the selection and utilization of medicines to treat self-recognized symptoms or ailments without consulting a physician [11]. It also includes the usage or re-usage of previously prescribed or unused drugs; direct purchasing of prescription drugs without consultation; and irrational use of over-the-counter medications [12].

The effects of self-medication includes antimicrobial resistance, drug interactions, addiction or dependence, and drug toxicity [13]. Potential risks of self-medication include incorrect self-diagnosis, adverse reactions, dangerous drug interactions, incorrect administration, incorrect dosage, incorrect therapy selection, masking of a severe disease, and the risk of drug dependence and abuse [14, 15]. Although the prevalence and correlates of self-medication have been widely discussed in many countries [16], there is limited data on the prevalence and factors associated with self-medication in underserved populations in informal settlements. We estimated the prevalence, and factors associated with self-medication for COVID-19 amongst urban slum dwellers in Jinja City.

## Methods

### Study design

This was a cross-sectional study among adult urban slum dwellers aged 18 years and above who reported having had confirmed or self-reported COVID-19 during the study period from January to December 2021.

### Study Area

This study was carried out in Jinja city in Eastern Uganda, a region with an estimated population of 331,079 people [17]. Jinja City has eight slum but this study was conducted in five randomly selected slums. The city also houses several educational institutions, commercial centres, industries, and factories, hence a growing sprawl of unplanned settlements. This has given rise to several slum communities around the city, which, among others, face a myriad of challenges, including poverty, poor health service delivery, and illiteracy. Slum dwellers sought treatment from drug shops or stores, public hospitals [18], local pharmacy [19], road-side herbalists [20] and traditional healers [20]. In Uganda, health care is provided in both private and public health centres. Uganda provides primary health care through its National Minimum Health Care Package at the lowest level health centres, while tertiary care is provided at the highest level health centres [21]. The largest public health facility in Jinja district is Jinja Regional Referral Hospital, which provides tertiary services, while several lower-level health facilities serve the majority of the population.

### Study population

Participants who were non-residents, and adults not capable of giving consent for any reason (mentally ill, critically sick, and those not able to respond), were excluded from the study as they did not represent the true residents of the slums.

### Sample size determination

The minimum sample size was estimated using the Kish Leslie sample size estimation formula for cross-sectional studies [22]. In this study, we used a self-medication prevalence of 72% previously reported in Namuwongo, a slum in Kampala [23] to estimate a sample size of 314. Through multistage sampling, the estimated sample size was adjusted with a design effect of 1.5. Therefore, with a sample size of 518 respondents, this study had 80% power to determine the prevalence of self-medication at 95% confidence interval.

### Sampling Procedures and participant recruitment

A multi-stage sampling strategy was employed to select study respondents. Five slums were randomly selected for this study out of eight slums in the study area, three slums were randomly selected in Southern Division and two slums in Northern Division.

In the first stage, the researchers used a stratified sampling method in which five of Jinja’s eight slums were randomly selected and proportionate to their size. In the second stage, the researcher selected households from the five selected slums. Later, a systematic sampling technique was further used, where every 41^st^ household (derived when we divided 21,284 total households by the 518-sample size calculated) was selected to reach the desired number of respondents per slum. This was done with the guidance of the local council leaders and/or Village Health Team (VHT) members. The local council or VHT members assisted in guiding the researchers to locate the boundaries of each slum, provided a list of households in each slum, and introduced the researchers to the community and the selected households. For the third stage, simple random sampling was used to select a participant in each household where more than one person met the inclusion criteria. The Kish Grid was used to select the required participant to avoid selection bias [24, 25]. Where there was no eligible person, the next household with an eligible respondent was selected. This was continued until the desired number of respondents of 518 was reached.

### Data collection methods and tools

A questionnaire was used to obtain quantitative data from the study respondents. The questionnaire used was adapted from previous studies [26-28] and modified to fit our study. The tool was translated into the local ‘*Lusoga*’ by language expert and pre-tested in Polota slum, which is similar to the Jinja city slums. The Cronbach’s Alpha coefficient reliability test was used to assess the data collection tool’s validity (coefficient alpha=0.83). The questionnaire was then uploaded onto KoBo Toolbox tool version 2022.1.2, a mobile phone application for data collection [29]. Data were collected by research assistants who were conversant with both the local language and English. Study respondents received consent from all respondents prior to the interviews. Data were collected using semi-structured questions installed on the KoBo Toolbox application tool version 2022.1.2.

Participants were asked (Yes/No) if they used any medications (medicines and herbs) for self-medication during COVID illness without a prescription or consultation with a medical practitioner/physician. Participants were also interviewed on several demographic and social characteristics. The characteristics were categorised as either predisposing, enabling, or need factors. Predisposing factors included: age, sex, ethnicity, occupation, education level, cultural beliefs, and societal influences. Enabling factors refer to availability constraints and included household income, medical insurance, proximity to medical services, advertisements, sources of advice, and legislation. Need factors are those which trigger the action to self-medicate, such as type of illness, duration of illness, the severity of illness, history of illness, and health status.

### Data management and analysis

The data collected were transferred into Microsoft Excel software and then STATA version 14.0 for further statistical analysis. Data cleaning was done by checking for variables with missing data and duplicates as well as transforming variables from string to numerical for analysis purposes to come up with a complete dataset. Outliers were identified using geographical methods and a Q–Q plot (quantile-quantile plot). The prevalence of self-medication for COVID-19 and symptoms suffered by slum dwellers was estimated.

### Statistical analysis

The factors associated with self-medication for COVID-19 were determined using Poisson regression with robust standard errors. We determined the crude prevalence rations (cPR) and adjusted prevalence ratios (APR) and 95% confidence intervals (CIs) for several covariates associated with self-medication for COVID-19. The backward elimination method was used and variables with a P-value ≤0.20 were considered at multivariable stage modelling stage. Only independent variables with a p-value≤ 0.05 were considered significant. All estimates were reported with a 95% confidence interval. Multi-collinearity was checked using scatter plots and no variables had correlation coefficient greater than 0.4. The model with the least Akaike information criterion (AIC) and Bayesian information criterion (BIC) values was selected as the final model. The goodness of fit test was used to test the fitness of the final model.

### Ethical Considerations

We obtained ethical approval to conduct this study from Makerere University, School of Public Health Higher Degrees Research and Ethics Committee (HDREC), approval number FWA00011353. Administrative clearance was also obtained from the Jinja city authorities. Written informed consent was obtained from all respondents before the interview. Confidentiality was maintained throughout the study.

## Results

### Characteristics of study participants

A total of 517 respondents were interviewed representing 99.80% response rate. The median age of respondents (IQR) was 31 (26-40) years and 40.42% (209/517) were within the age group of 30-39. In addition, 58.99% (305/517) were males. Among the respondents, 52.80% (273/517) had attained secondary-level education, 71.76% (371/517) were married while 50.68% (262/517) had a family monthly income of less than 100,000 Uganda shillings (approximately ≤ 26.32 U.S. dollars), median income (IQR) being 100,000 (50,000-300,000) **Table 1**.

**Table 1:** Socio-demographic information of the respondents.

| Variable | Frequency (n=517) | Percentage (%) |
| --- | --- | --- |
| **Age (years)** |  |  |
| 20-29 | 125 | 24.18 |
| 30-39 | 209 | 40.43 |
| 40-49 | 105 | 20.31 |
| ≥50 | 78 | 15.09 |
| Median age (IQR) years | 31 (26-40) |  |
| **Sex** |  |  |
| Male | 305 | 59.99 |
| Female | 212 | 41.01 |
| **Religion** |  |  |
| Anglican | 112 | 21.66 |
| Catholic | 135 | 26.11 |
| Muslim | 159 | 30.75 |
| Pentecostal | 90 | 17.41 |
| Other | 21 | 4.06 |
| **Education** |  |  |
| No formal | 47 | 9.09 |
| Primary | 137 | 26.50 |
| Secondary | 273 | 52.80 |
| Tertiary | 60 | 11.61 |
| **Marital Status** |  |  |
| Never Married | 97 | 18.76 |
| Married | 371 | 71.76 |
| Separated | 26 | 5.03 |
| Widower/widow | 23 | 4.45 |
| **Monthly Income (UGX)** |  |  |
| <100, 000 | 262 | 50.68 |
| 100,001-200,000 | 89 | 17.21 |
| 200,001-300,000 | 68 | 13.15 |
| 300,001-400,000 | 34 | 6.58 |
| >400,000 | 64 | 12.38 |
| Median income (IQR) | 100,000 (50,000-300,000) |  |

**IQR:** Inter-quartile range

### Prevalence of self-medication for COVID-19

Overall, the prevalence of self-medication for COVID-19 was 87.23% (451/517), 95% CI: [84-90] among slum dwellers in Jinja. Of the 451 respondents who self-medicated for COVID-19, 56.76% (256/451) agreed that they were aware of the risks associated with self-medication, while 15.74% (71/451) disagreed. The most common symptoms suffered were difficulty in breathing, sore throat, loss of appetite, chest pain, fever and other (dizziness, vomiting, bloating of the stomach and hearing loss) symptoms. **Fig 1 Symptoms of COVID-19 among respondents who self-medicated**

### Factors associated with self-medication for COVID-19

After controlling for potential confounders, being old (>50 years) [APR: 1.12, 95% CI:1.05, 1.20], female sex [APR: 1.07, 95% CI: 1.02, 1.13], symptoms severity: minor symptoms [APR: 1.62, 95% CI: 1.25, 2.11]; severe symptoms [APR: 1.51, 95% CI: 1.16, 1.96], having medical insurance [APR: 0.63, 95% CI: 0.46, 0.87], access to internet [APR: 1.13, 95% CI: 1.07, 1.20], family/friends influence [APR: 1.27, 95% CI: 1.12, 1.42], and anxiety/fear of being quarantined [APR: 1.35, 95% CI: 1.14, 1.60] were significantly associated with higher rates of self-medication for COVID-19. Being aware of laws against self-medication was also found to be associated with a lower risk of self-medication [APR: 0.89, 95% CI: 0.81, 0.97]. On the other hand, having had a prior COVID-19 test, previous experience with the disease, health facility being far, long waiting time at the health facility, short duration of the disease or its symptoms, poor present health status, highest education levels, marital status and household monthly income were all found not statistically significate. **Table 2**

**Table 2:** Bivariate and multivariable factors associated with self-medication among slum dwellers in Jinja City.

| Variables | Have you ever self-medicated<br>(n=517) |  | CPR (95% CI)* | APR (95%CI)** | p-value |
| --- | --- | --- | --- | --- | --- |
|  | No n (%)<br>n=66 | Yes n (%)<br>n=451 |  |  |  |
| **Sex** |  |  |  |  |  |
| Male | 37 (17.45) | 175 (82.55) | Ref | Ref |  |
| Female | 29 (9.51) | 276 (90.49) | 1.07 (1.02, 1.13) | 1.1 (1.0, 1.1) | **0.006** |
| **Age (years)** |  |  |  |  |  |
| 20-29 | 19(15.20) | 106(84.80) | Ref | Ref |  |
| 30-39 | 29(13.88) | 180(86.12) | 1.02(0.93, 1.11) | 1.03(0.97, 1.09) | 0.384 |
| 40-49 | 13(12.38) | 92(87.62) | 1.03(0.93, 1.15) | 1.05(0.98, 1.12) | 0.133 |
| 50+ | 5(6.41) | 73(93.59) | 1.10(1.00, 1.21) | 1.12(1.05, 1.20) | **0.001** |
| **Anxiety and or fear of quarantine** |  |  |  |  |  |
| No | 21(42.86) | 28(57.14) | Ref | Ref |  |
| Yes | 9(2.82) | 310(97.18) | 1.70(1.33, 2.17) | 1.35(1.14, 1.60) | **<0.001** |
| **Have access to medical insurance** |  |  |  |  |  |
| No | 46(9.52) | 437(90.48) | Ref | Ref |  |
| Yes | 20(58.82) | 14(41.18) | 0.46(0.30, 0.68) | 0.63(0.46, 0.87) | **0.005** |
| **Internet/ advertisement** |  |  |  |  |  |
| No | 47(21.17) | 175(78.83) | Ref | Ref |  |
| Yes | 18(6.12) | 276(93.88) | 1.19(1.11, 1.28) | 1.13(1.07, 1.20) | **0.001** |
| **Influenced by my family or friends** |  |  |  |  |  |
| No | 48(46.15) | 56(53.85) | Ref | Ref |  |
| Yes | 18(4.36) | 395(95.64) | 1.78(1.48, 2.13) | 1.27(1.12, 1.42) | **<0.001** |
| **Aware of any law on medicine use** |  |  |  |  |  |
| No | 39(9.11) | 389(90.89) | Ref | Ref |  |
| Yes | 27(30.34) | 62(69.66) | 0.77(0.67, 0.88) | 0.89(0.81, 0.97) | **0.008** |
| **Symptom severity** |  |  |  |  |  |
| No symptoms | 39 (67.24) | 19 (32.76) | Ref | Ref |  |
| Minor symptoms | 11(5.14) | 203(94.86) | 2.89(2.00, 4.19) | 1.62(1.25, 2.11) | **<0.001** |
| Severe symptoms | 16(6.53) | 229(93.47) | 2.85(1.97, 4.13) | 1.51(1.16, 1.96) | **0.002** |
| **Prior COVID-19 test** |  |  |  |  |  |
| No | 22(7.12) | 287(92.88) | Ref | Ref |  |
| Yes | 18(11.69) | 136(88.31) | 0.95(0.89,1.01) | 1.01(0.96,1.06) | 0.736 |
| **Previous experience with the disease** |  |  |  |  |  |
| No | 49(11.53) | 376(88.47) | Ref | Ref |  |
| Yes | 17(18.48) | 75(81.52) | 0.92(0.83, 1.02) | 0.96(0.89,1.03) | 0.236 |
| **The health facility is far** |  |  |  |  |  |
| No | 49(11.64) | 372(88.36) | Ref | Ref |  |
| Yes | 17(17.71) | 79(82.29) | 0.93(0.84, 1.03) | 0.93(0.85,1.01) | 0.091 |
| **Long waiting time at the health facility** |  |  |  |  |  |
| No | 45(14.61) | 263(85.39) | Ref | Ref |  |
| Yes | 21(10.05) | 188(89.95) | 1.05(0.99, 1.12) | 0.01(0.96,1.06) | 0.679 |
| **The short duration of the disease or its symptoms/signs** |  |  |  |  |  |
| No | 51(14.37) | 304(85.63) | Ref | Ref |  |
| Yes | 15(9.26) | 147(90.74) | 1.06(0.99, 1.13) | 1.02(0.97, 1.07) | 0.375 |
| **Poor present health status** |  |  |  |  |  |
| No | 41(17.23) | 197(82.77) | Ref | Ref |  |
| Yes | 25(8.96) | 254(91.04) | 1.10(1.03, 1.18) | 0.97(0.92, 1.02) | 0.214 |
| **Education** |  |  |  |  |  |
| No education | 5(10.64) | 42(89.36) | Ref | Ref |  |
| Primary | 15(10.95) | 122(89.05) | 1.00(0.89, 1.12) | 0.98(0.89, 1.08) | 0.733 |
| Secondary | 30(10.99) | 243(89.01) | 1.00(0.89, 1.12) | 1.03(0.93, 1.14) | 0.548 |
| Tertiary | 16(26.67) | 44(73.33) | 0.82(0.68, 0.98) | 1.00(0.87, 1.14) | 0.992 |
| **Marital Status** |  |  |  |  |  |
| Never Married | 20(20.62) | 77(79.38) | Ref | Ref |  |
| Married | 41(11.08) | 330(88.92) | 0.89(0.81, 0.99) | 1.00(0.93, 1.08) | 0.987 |
| Separated | 2(7.69) | 24(92.31) | 1.04(0.92, 1.17) | 0.97(0.85, 1.11) | 0.626 |
| Widower/widow | 3(13.04) | 20(87.98) | 0.98(0.83, 1.15) | 1.03(0.92, 1.15) | 0.605 |
| **Monthly income (UGX)<sup>¶</sup>** |  |  |  |  |  |
| 0-100,000 | 42(16.03) | 220(83.97) | Ref | Ref |  |
| 100001-200,000 | 8(8.99) | 81(91.01) | 1.08(0.99, 1.18) | 1.04(0.97, 1.11) | 0.245 |
| 200,001-300,000 | 8(11.76) | 60(88.24) | 1.05(0.95, 1.16) | 0.99(0.93, 1.05) | 0.700 |
| 300,001-400,000 | 3(8.82) | 31(91.17) | 1.09(0.97, 1.22) | 0.97(0.88, 1.06) | 0.504 |
| 400001-max | 5(7.81) | 59(92.19) | 1.10(1.00, 1.20) | 0.99(0.92, 1.06) | 0.781 |

**Note:** CPR= \*Crude Prevalence Ratio, APR= \*\*Adjusted Prevalence Ratio. <sup>¶</sup>1USD=3800 Uganda shillings (UGX)

## Discussion

Our findings show that there is a high prevalence of COVID-19 self-medication among urban slum dwellers. This was demonstrated by the fact that 87.2% of urban slum dwellers in Jinja city reported using self-medication. The high prevalence of self-medication may be a result of the challenges related to healthcare access brought about by the pandemic (high health costs, travel restrictions, and a shortage of health staff). Studies conducted in Bangladesh and Uganda found almost a similar prevalence to this current study of 88.3% [30] and 88% [31] respectively. Further, a study conducted among the general public found a decrease in self-medication practices during the COVID-19 pandemic lockdown compared to the practice before the lockdown [31]. This disparity could be explained by the fact that during the COVID-19 lockdown, the public was not moving freely, making it difficult for them to obtain drugs. The prevalence of self-medication in our study, however, was way higher than that reported among residents of urban slums [32]. This discrepancy can be explained by the fact that both of these studies were conducted way before the COVID-19 pandemic which started almost three years later.

Factors found to be associated with self-medication for COVID-19 included age, sex, anxiety and or fear of quarantine, access to medical insurance, internet/advertisement, awareness of the laws regarding medicine use, family/ friend influence, and symptom severity. Our study revealed that respondents who were 50 years and older showed a 12% increase in the likelihood of self-medication compared to those in the 20-29 age bracket. This could be explained by the higher COVID-19 mortality among the elderly, hence creating a higher perceived risk among this population. In addition, older people are prone to chronic infections, thus being susceptible to infections like COVID-19 [33]. In addition, studies conducted in Uganda, Eritrea, Ethiopia, Nigeria, Belgrade and Cameroon have reported that as one gets older, they acquire accumulated knowledge due to disease experience [33-40]. Contrary studies have been done in Jabalpur [32] and Syrian [41], where individuals aged 18 to 40 years were more likely to self-medicate. This age difference could be due to the population dynamics of the respondents.

Our study showed that females were two times more likely to self-medicate than males. The reason for the association between being a female and self-medication is not known, but in the context of the COVID-19 outbreak, greater anxiety among women, as described in Iran and Italy, cannot be excluded [42, 43]. In addition, this might have been due to the exacerbated stress among women associated with the COVID-19 pandemic and efforts to mitigate its spread [44]. This is consistent with results in studies conducted in Spain and elsewhere that found that more females self-medicated compared to their counterparts [45, 46]. However, there are controversial data on the relationship between sex and self-medication as studies conducted in Iran and the United Arab Emirates [47, 48] respectively did not report any association. This difference could be due to these studies having been conducted among different study populations.

In the current study, respondents who reported anxiety and fear of being quarantined had a 35% increase in the likelihood of self-medication compared to their counterparts. It is important to note that during the period under study, COVID-19-related morbidity and mortality were very high in Uganda and could have increased anxiety levels and fear of infection. Anxiety could have exacerbated self-medication as people tried their level best to prevent the infection, treat any present signs and symptoms, as well as boost immunity to fight off disease. In addition, the mass fear of COVID-19, termed “corona phobia” [49-51] coupled with stringent infection prevention and control measures, was reported to have caused a lot of depression, confusion, fear, anger, and despair among people. This consequently resulted in substance use, which is self-medication [49]. Similar findings have been reported by other scholars [52-54].

This study found that having access to medical insurance was more protective compared to those who did not have medical insurance. At the peak of the second wave of COVID-19 in Uganda, the public healthcare system could not ably serve all confirmed and suspected COVID-19 cases effectively, since treatment was expensive in private facilities, people who had insurance accessed treatment easily in these private facilities, and those offered better services [55, 56]. The majority of those who accessed private facilities were likely to have medical insurance, as supported by findings of a study conducted in Iran [57].

The findings of this study show that individuals who reported access to the internet and advertisements showed a 13% increase in the likelihood of self-medication compared with those who did not. Comparable studies have been conducted in Iran and India [58, 59]. There was infodemia about the COVID-19 pandemic due to much information in both digital and physical environments. This resulted in confusion and risk-taking behaviours that could harm health, foster distrust in health authorities or systems, and undermine the public health response, leading to self-medication [60, 61]. This information was used by the public as a replacement for medical consultations [59, 62]. In this response, the WHO designed a taxonomy to address infodemia, which was piloted in individual countries and WHO regions to generate localized insights and actions [60]. Furthermore, these findings indicated that the likelihood of self-medication for COVID-19 increased by 27% due to influence from family and friends. This finding reflects the familial clustering of the disease by the respondents since some reported self-medication for the prevention of COVID-19 infection. A possible reason is that family and friends tend to live together and often communicate with each other. Therefore, individuals who tested positive for COVID-19 might have taken the initiative to inform their recent acquaintances, who, under the scare of acquiring infection, resorted to self-medication in the hope of prevention [62-66].

Respondents who experienced minor and severe symptoms had a higher likelihood of 62% and 51%, respectively, of self-medicating compared to those who had no symptoms. These results mean that there was a high likelihood of self-medication among slum dwellers who either experienced minor or severe symptoms. Individuals with minor symptoms ignored the disease and thought that since it was not severe, they would easily recover in a short time. This is supported by studies conducted in Nigeria, and Ethiopia and a systematic review [36, 38, 67] respectively. On the other hand, those with severe symptoms viewed themselves as having the severe form of the disease that requires immediate attention and cannot be delayed due to issues of health facility access, affordability, drug stock-outs, and long waiting times. This is comparable to studies conducted in Brazil [68] and the USA [69]. Respondents who knew laws regarding medicine use were more likely to self-medicate, which is in line with a study conducted in Colombia [70]. During the COVID-19 era, the greater access of people to the media, the internet, and their ability to understand information about the medicine and its related laws on use found on social networks may explain this finding. Since many patients get knowledge about drugs from previous prescriptions [71], physicians should limit superfluous prescriptions of antibiotics and implement guideline-based practices.

### Limitations of the study

The study had potential limitation. First, this study based on self-reporting, hence more likely to lead to social desirability bias because of stigma, anxiety, and fear. However, this was minimized by assuring the respondents before the study about information confidentiality, reviewing respondents’ medical records while conducting data collection, and checking the Jinja City health database on COVID-19 test results. This study was conducted during the COVID-19 pandemic when people had fear and stigma about freely interacting with anyone for fear of contracting the disease. However, this was addressed by ensuring all the research assistants observed COVID-19 SOPs while in the field. These findings can be used to plan and manage future epidemics or any emergencies of public health concern and also guide the engagement of slum dwellers towards a reduction in the prevalence of self-medication. Therefore, these results could be generalizable to the general population of urban slum dwellers.

## Conclusion

This study reports a very high prevalence of self-medication for COVID-19 among urban slum dwellers in Jinja City. This is a signal for potential future antimicrobial resistance outbreaks, especially in vulnerable groups like urban slum dwellers. Self-medication has been known to result in unintended healthcare outcomes such as antimicrobial resistance, adverse effects, drug interactions, and drug dependence, among others. According to WHO, the COVID-19 pandemic situation is likely to be prolonged for years and will have a huge effect on the social-economic and psychosocial impact on people’s lifestyles and behaviour. This poses a more serious threat that might push people to self-medicate. Therefore, MoH and NDA should design stringent measures for the prescription, sale, and use of medicine, not forgetting to regulate advertisements in addition to sensitization of the slum dwellers.

## Recommendations

Based on the findings, the researchers suggest the following recommendations to the Ugandan government and other stakeholders. The Ministry of Health needs to adopt WHO-approved guidance on the detection of info-demic signals and implement medical insurance policy to benefit the most vulnerable populations like slum dwellers. The National Drugs Authority (NDA) should put in place stringent mechanisms to regulate the prescription, sale, hawking, and use of conventional medicines, especially antibiotics. Also, NDA should liaise with herbal medicine practitioners to establish active elements in socially acceptable and effective herbal medicines. The City Health Team (CHT) should control and manage the sale of any medicine by conducting routine inspections, routine support supervision of all sellers of medicines and mass sensitisation of all stakeholders. During health crises, such as the COVID-19 pandemic, slum dwellers and the general public ought to practice good health-seeking behaviours.

## Data Availability

Data Access / Ethics Committee (contact via) for researchers who meet the criteria for access to confidential data.

## Notes

### Competing Interest Statement

The authors have declared that no competing interests exist.

### Funding Statement

The authors received no specific funding for this work.

### Author Declarations

We obtained ethical approval to conduct this study from Makerere University, School of Public Health Higher Degrees Research and Ethics Committee (HDREC), approval number FWA00011353.

